# TRENDS-Thai: decadal trends of dengue, chikungunya, and hand, foot, and mouth disease in Thailand (2016–2025): a multi-disease time-series analysis of COVID-19 disruption

**DOI:** 10.64898/2026.05.21.26353796

**Authors:** Wannarat Pongpirul, Mohamed Mustaf Ahmed, Krit Pongpirul

## Abstract

**Introduction:** Dengue, chikungunya, and hand, foot, and mouth disease (HFMD) are priority notifiable infections in Thailand. Whether vector-borne and contact-mediated diseases responded differently to the coronavirus disease 2019 pandemic has not been quantified within a unified national surveillance framework over an extended period.

**Methods:** We conducted an ecological interrupted time-series analysis using weekly province-level notifiable disease surveillance data from epidemiological week 1 of 2016 to week 53 of 2025 across all 77 Thai provinces. Incidence per 100,000 population was calculated using year-specific civil registration population denominators. Segmented quasi-Poisson regression with two Fourier harmonics for annual seasonality was fitted, with the primary pandemic onset defined as week 1 of 2020 and two alternative onset definitions prespecified for sensitivity analysis.

**Results:** The analysis included 40,579 province-week observations across 527 epidemiological weeks, comprising 790,263 dengue, 32,265 chikungunya, and 713,822 HFMD cases nationally. Immediate incidence rate ratios at pandemic onset were 0.39, 0.54, and 0.51 for dengue, chikungunya, and HFMD, respectively. Sustained post-onset trends diverged across diseases, with declining trajectories for the two vector-borne infections and a positive post-onset slope for hand, foot, and mouth disease. Dengue rebounded above pre-pandemic levels by 2023, chikungunya remained quiescent through 2025, and HFMD exceeded its pre-pandemic baseline by approximately 26%.

**Conclusion:** Vector-borne and contact-mediated diseases in Thailand followed sharply contrasting decadal trajectories that tracked the transmission ecologies of each pathogen. These findings support transmission-mode-specific pandemic-resilient surveillance, accelerated arboviral and enteroviral vaccine deployment, and integrated vector management.

## Introduction

Population-level shocks rarely affect all infectious diseases equally. Vector-borne and contact-mediated infections follow fundamentally different transmission dynamics and may therefore respond differently to disruptions in human behavior, mobility, and surveillance. The coronavirus disease 2019 (COVID-19) pandemic produced a synchronous, multi-domain shock: large-scale movement restrictions and non-pharmaceutical interventions sharply reduced population mixing in affected regions ^1^, coinciding with sustained suppression of seasonal influenza and other respiratory viruses during 2020-2021 ^2^, and disrupted routine health programs in low- and middle-income countries to a degree projected to translate into substantial excess maternal and child mortality ^3^. The pandemic therefore provides an unusual natural experiment to examine how diseases with contrasting transmission ecologies respond to a common external disruption within a unified surveillance framework.

Three priority notifiable infections in Thailand—dengue, chikungunya, and hand, foot, and mouth disease (HFMD)—represent the two principal transmission mechanisms relevant to this question. Geospatial modelling suggests that approximately half of the world’s population resides in areas suitable for dengue transmission, with the highest suitability concentrated across South and Southeast Asia ^4^. Global burden analyses estimate that incident dengue episodes nearly doubled from approximately 30.7 million in 1990 to 56.9 million in 2019 ^5^. Dengue has continued to expand globally as *Aedes*-mediated transmission extends into new tropical and subtropical environments ^6^, and Thailand remains hyperendemic with recurrent province-level outbreaks and spatially clustered epidemic dynamics ^7^. Thailand also experienced a major nationwide chikungunya outbreak during 2018-2019, driven by the East/Central/South African Indian Ocean lineage ^8^. HFMD, predominantly caused by enterovirus A71 (EV-A71) and coxsackieviruses A6 (CV-A6) and A16 (CV-A16), represents one of the leading pediatric viral syndromes in the Western Pacific region ^9^. Thai molecular surveillance has documented sequential EV-A71 subgenogroup turnover and the emergence of CV-A6 as a dominant outbreak driver during the early 2010s ^10^.

Single-disease studies have described pandemic-era changes in each disease independently and within disease-specific analytic frameworks. Thai dengue notifications declined sharply during the early pandemic before rebounding in subsequent years; over a longer pre-pandemic horizon, the mean age of reported dengue cases had already been increasing ^11^. Chikungunya activity remained relatively quiescent following the 2018-2019 outbreak, whereas HFMD exhibited a marked pandemic-era decline followed by an above-baseline rebound. However, the differential responses of vector-borne and contact-mediated diseases have not been quantified within a harmonized multi-disease framework spanning a full decade. Likewise, the extent of geographic heterogeneity across a national surveillance system and whether post-pandemic trajectories align more closely with transmission ecology than disease-specific epidemic history remain unclear.

Distinguishing pandemic-associated changes from underlying secular trends and seasonal cycles requires harmonized multi-disease surveillance data on a consistent temporal grid, together with an analytical framework capable of separating immediate level changes at the pandemic onset from sustained post-onset trend changes. Segmented regression provides an established approach for estimating these components at a defined intervention point ^12^. This framework can be adapted for count surveillance data using quasi-Poisson estimation with Fourier seasonality terms and population offsets. Sensitivity analyses using alternative pandemic onset definitions, combined with overdispersion-adjusted variance estimation, reduce dependence on arbitrary intervention timing and seasonal misspecification—two major internal validity threats in interrupted time-series designs.

This study used ten years (2016-2025) of weekly province-level notifiable disease surveillance data from all 77 provinces in Thailand to (i) characterize decadal trajectories of dengue, chikungunya, and HFMD within a unified national framework; (ii) quantify immediate and sustained changes in incidence associated with the onset of the COVID-19 pandemic, with formal sensitivity analyses for alternative onset definitions; and (iii) compare the responses of vector-borne and contact-mediated diseases to a common external disruption to evaluate whether observed patterns are more consistent with underlying transmission ecology than disease-specific epidemic cycles.

## Methods

### Study design and setting

We conducted a national ecological interrupted time-series analysis in accordance with the Strengthening the Reporting of Observational Studies in Epidemiology (STROBE) guidelines ^13^. The completed checklist is provided in Supplementary File S1. The study setting was Thailand and included all 77 provinces over the period from 2016 to 2025. The COVID-19 pandemic was conceptualized as a population-level external disruption affecting three notifiable diseases with distinct transmission pathways within a shared surveillance system. Because the analysis used the complete national passive surveillance census rather than sampled observations, no formal sample size calculation was performed.

### Data sources, eligibility, and measurement

All weekly province-level notifications for dengue fever (International Classification of Diseases, Tenth Revision (ICD-10) code A90), dengue hemorrhagic fever (A91), dengue shock syndrome (A91.1), chikungunya (A92.0), and HFMD (B08.4, B08.5) reported between January 1, 2016, and December 31, 2025, were eligible for inclusion.

Case counts were obtained from the Ministry of Public Health (MOPH) 506 notifiable disease surveillance system, which receives mandatory notifications from public and private health facilities across all provinces in Thailand.

Year-specific provincial population denominators were obtained from the Department of Provincial Administration (DOPA) Bureau of Registration Administration. Province polygons used for spatial analyses were obtained from the Royal Thai Survey Department and distributed through the United Nations Office for the Coordination of Humanitarian Affairs (UNOCHA) Common Operational Dataset for Thailand administrative boundaries (ADM-1).

The harmonised province-week records used in this analysis constitute the Thailand release of TRENDS (Temporal Recent Epidemiology of Notifiable Diseases in Southeast Asia) and have been deposited in a public repository to promote transparency, reproducibility, and reuse in epidemiological research (TRENDS-Thai Dataset, Zenodo: The harmonised province–week records used in this analysis constitute the Thailand release of **TRENDS (Temporal Recent Epidemiology of Notifiable Diseases in Southeast Asia)** and have been deposited in a public repository to promote transparency, reproducibility, and reuse in epidemiological research (TRENDS-THAI Dataset, Zenodo: https://zenodo.org/records/20269896)^14^.

The final analytical dataset contained 40,579 province-week observations across 527 epidemiological weeks. Completeness across the province x epidemiological week grid was 100%; therefore, no imputation procedures were required.

### Variables and outcome definitions

The primary outcome was the weekly incidence per 100,000 population:

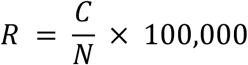

where *R* denotes weekly incidence per 100,000 population, *C* is the reported weekly case count, and *N* is the year-specific provincial population.

Only reported cases were analyzed. Individual-level characteristics were unavailable, and serotype/genotype information was not incorporated.

A composite dengue series was defined as the sum of the three ICD-10 severity categories because all represent *Aedes*-mediated infection and classification boundaries may vary with clinical practice.

Chikungunya and HFMD were analyzed separately.

Diseases were grouped a priori into two transmission categories for the mechanism-level interpretation: vector-borne (dengue and chikungunya) and contact-mediated (HFMD).

The exposure variable was the pandemic period. Three descriptive phases were predefined: pre-COVID (2016–2019), pandemic (2020–2022), and post-COVID (2023–2025). For interrupted time-series modelling, the primary pandemic onset was defined as epidemiological week 1 of 2020. Two alternative onsets were prespecified for sensitivity analysis: week 12 of 2020 (first national lockdown) and week 42 of 2020 (second epidemic wave).

### Statistical analysis

Descriptive analyses included 8-week centered rolling mean of the national weekly incidence per 100,000 across the full window, mean monthly incidence by pandemic phase, province-level cumulative incidence per 100,000 person-years, and percent change in province-level annual incidence relative to the pre-COVID phase.

Segmented quasi-Poisson regression was fitted to national weekly counts to estimate immediate level change and sustained post-onset slope change, following established interrupted time-series methodology ^15^. The model was specified as:

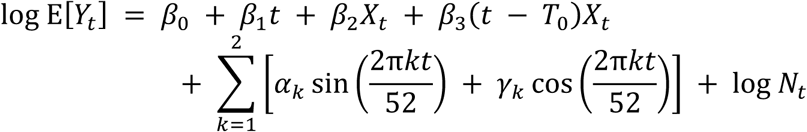

where *Yₜ* denotes national weekly case count; *t*, time in weeks, *Xₜ*, pandemic indicator; *T₀*, pandemic onset, *β₀* to *β₃* are the segmented regression coefficients, αₖ and γₖ are Fourier harmonic coefficients for annual seasonality with a period of 52 weeks, and log Nₜ is the offset for the year-specific national population. A quasi-Poisson family was used to accommodate the overdispersion in the count series. Coefficients were exponentiated to incidence rate ratios (IRR) with 95% confidence interval (CI). The post-onset slope was re-expressed as an annualized % change, as follows:

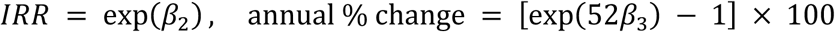

A no-COVID counterfactual trajectory was generated by fitting the seasonal baseline model to pre-pandemic weeks only and projecting forward; the chikungunya counterfactual was fit on 2016 to 2017 data because the 2018 to 2019 outbreak distorted the pre-pandemic baseline. Pairwise temporal coupling between diseases was assessed using the cross-correlation of de-seasonalised national weekly log rates at lags from minus 26 to plus 26 weeks, with the 95% null band defined as plus or minus 1.96 divided by the square root of the sample size. To compare diseases on a single baseline-anchored axis, vector-borne and contact-mediated weekly rates were indexed to their own pre-COVID weekly mean set to 100, as follows:

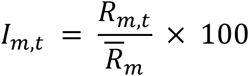

where m denotes the transmission mode (vector-borne for dengue and chikungunya, contact-mediated for HFMD), Rₘ,ₜ is the weekly rate for mode m at week t, and Rmₘ is the pre-COVID weekly mean rate for mode m. Two-sided p-values less than 0.05 were considered statistically significant.

### Bias and sensitivity analyses

Several measures were implemented to reduce bias. Outcome misclassification was minimized by using standardized ICD-10 case definitions within the MOPH 506 surveillance system. Passive surveillance may be affected by underreporting and cannot be fully corrected. Because under-ascertainment during COVID was expected to operate primarily through temporal disruption rather than systematic disease-specific bias, robustness was evaluated using: alternative pandemic onset definitions, explicit time adjustment, and overdispersion-adjusted estimation. Long-term trends and seasonality was addressed through continuous time adjustment and Fourier harmonics.

Prespecified subgroup analyses were conducted: by disease (separate models for dengue, chikungunya, and HFMD) and by transmission mode (vector-borne versus contact-mediated). Sensitivity analyses re-estimated interrupted time-series models under each alternative onset specification. All analyses were performed using Python 3.13.

### Ethics approval

This study used aggregated, de-identified surveillance data reported through the national notifiable disease surveillance system of Thailand and did not involve contact with human participants or access to identifiable individual-level information.

The study protocol was reviewed by the Institutional Review Board of Bamrasnaradura Infectious Diseases Institute (BIDI) and was determined to meet criteria for exemption from full ethical review as minimal-risk secondary research using aggregated surveillance data (IRB Reference: S002h/69_Exempt).

Permission to use surveillance data was obtained through institutional and administrative processes of the Ministry of Public Health, Thailand. All analyses were conducted in accordance with applicable ethical standards and data governance requirements. Public dissemination of the accompanying dataset, if applicable, includes only province-level aggregated data and contains no personal identifiers or individual-level records.

## Results

### Study cohort and disease groupings

The analysis included 40,579 province-week observations across 77 Thai provinces and 527 epidemiological weeks from 2016 to 2025. During the study period, 790,263 dengue, 32,265 chikungunya, and 713,822 HFMD cases were reported nationally. For mechanism-level comparison, dengue and chikungunya was classified as vector-borne diseases transmitted by *Aedes* mosquitoes, whereas HFMD was classified as a contact-mediated disease predominantly affecting children younger than five years (Table 1).

**Table 1.** Disease groupings used in the analysis.

| Disease | ICD-10 codes | Transmission category |
| --- | --- | --- |
| Dengue | A90, A91, A91.1 | Vector-borne (Aedes aegypti and Aedes albopictus) |
| Chikungunya | A92.0 | Vector-borne (Aedes aegypti and Aedes albopictus) |
| Hand, foot, and mouth disease | B08.4, B08.5 | Contact-mediated (paediatric, predominantly under five years) |

Note: ICD-10, International Classification of Diseases, 10th revision

### Annual case burden and population denominator

Substantial interannual variation was observed across all three diseases over the study period. Dengue exhibited the greatest fluctuation, decreasing to 10,524 reported cases in 2021 during the pandemic period before increasing to a post-pandemic peak of 160,841 cases in 2023. Chikungunya displayed a distinct epidemic profile, with a nationwide surge during 2018-2019 culminating in 11,721 cases in 2019, followed by persistently low activity thereafter. HFMD declined sharply during the pandemic period, reaching 19,007 cases in 2021, before rebounding to 115,264 cases by 2025 (Table 2).

**Table 2.** Annual case counts and national population, 2016 to 2025.

| Year | Dengue cases | Chikungunya cases | HFMD cases | National population |
| --- | --- | --- | --- | --- |
| 2016 | 64,174 | 18 | 80,348 | 65,931,550 |
| 2017 | 53,961 | 10 | 70,733 | 66,188,503 |
| 2018 | 87,656 | 3,581 | 70,191 | 66,413,979 |
| 2019 | 129,757 | 11,721 | 67,738 | 66,558,935 |
| 2020 | 72,557 | 11,334 | 33,805 | 66,186,727 |
| 2021 | 10,524 | 661 | 19,007 | 66,171,439 |
| 2022 | 46,655 | 1,309 | 100,140 | 66,090,475 |
| 2023 | 160,841 | 1,486 | 65,673 | 66,052,615 |
| 2024 | 105,986 | 740 | 90,923 | 65,951,210 |
| 2025 | 58,152 | 1,405 | 115,264 | 65,809,011 |

Note: Case counts are notified cases reported to the national surveillance system. National population is the mid-year estimate

The national population denominator remained relatively stable throughtout the study period, increasing from 65.9 million in 2016 to a peak of 66.6 million in 2019, followed by a gradual decline to 65.8 million in 2025. Annual incidence estimates were therefore interpreted primarily as reflecting changes in disease occurrence rather than major shifts in population size.

### National weekly trajectory of incidence

The 8-week centered rolling mean of national weekly incidence per 100,000 population demonstrated pronounced annual seasonality across all three diseases during 2016-2025. Dengue exhibited the largest temporal fluctuation, with the highest smoothed incidence occurring in August 2023 following substantial suppression during the pandemic period. Chikungunya showed a distinct epidemic profile, reaching its highest smoothed incidence in July 2020, consistent with the tail of the nationwide 2018-2019 outbreak. HFMD reached its highest smoothed incidence in August 2022, reflecting a rapid rebound after marked reductions during the pandemic years. Across all three diseases, prolonged troughs were observed during the pandemic period, followed by recovery to, or in some instances exceeding, pre-pandemic levels during the post-COVID phase (Figure 1).

**Figure 1.**
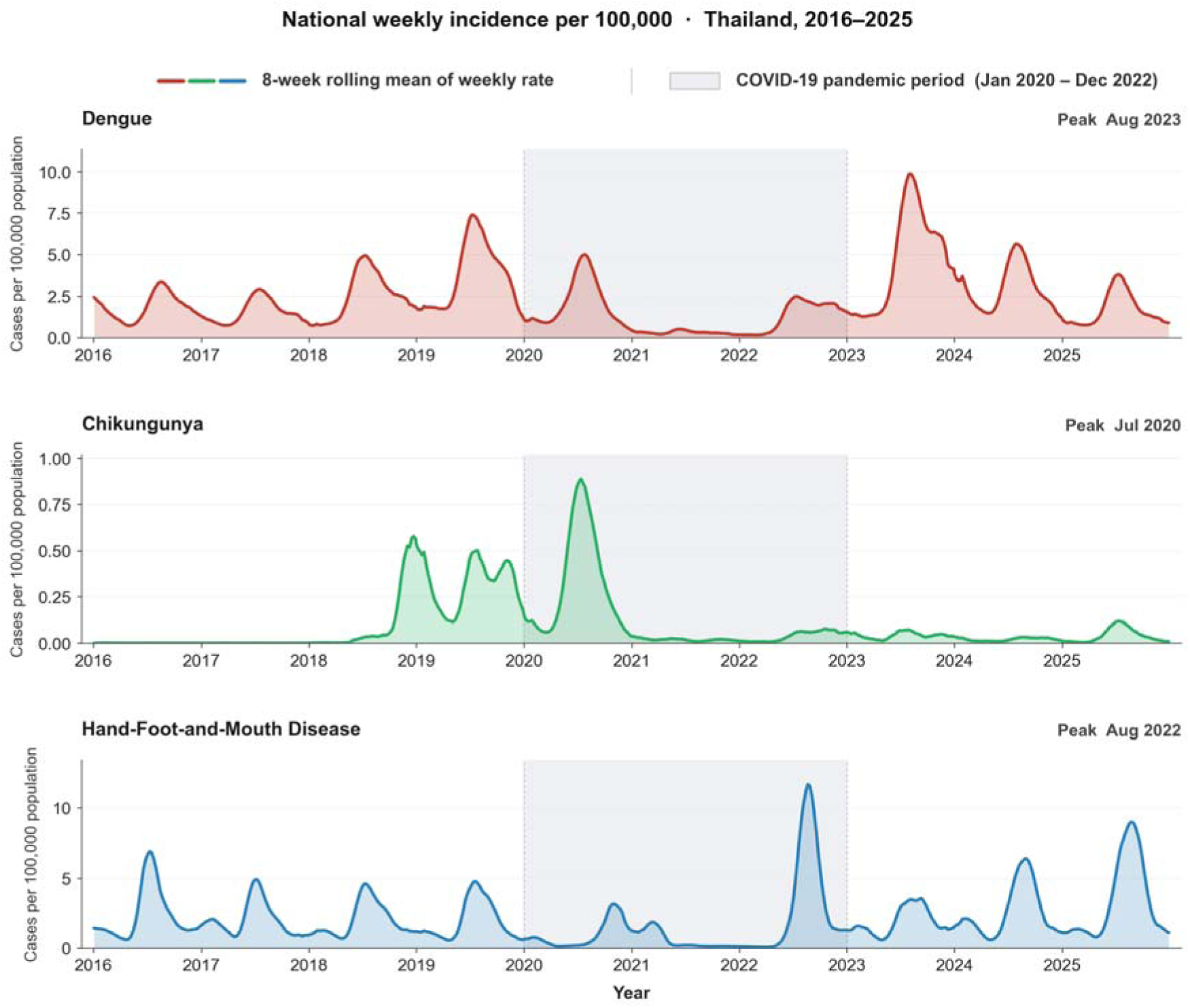
Smoothed national weekly incidence per 100,000 population for dengue, chikungunya, and hand, foot, and mouth disease in Thailand, 2016 to 2025.

### Within-dengue severity composition

Dengue notifications were further examined according to the three clinical severity strata. Although all strata broadly followed the temporal pattern of total dengue incidence across the pre-COVID, pandemic, and post-COVID periods, their relative composition changed progressively over the decade. The proportion of dengue fever (DF) increased from approximately 60% in 2016 to 73%–75% during the pandemic and post-COVID periods, whereas the proportion of dengue hemorrhagic fever (DHF) declined correspondingly from approximately 39% to 24%–29%. Dengue shock syndrome (DSS) remained consistently uncommon, accounting for approximately 1% of reported cases throughout the study period. The shift toward a greater proportion of DF and a lower proportion of DHF began before the pandemic and continued into the post-COVID period, suggesting that the observed change was not limited to the pandemic period alone (Figure 2).

**Figure 2.**
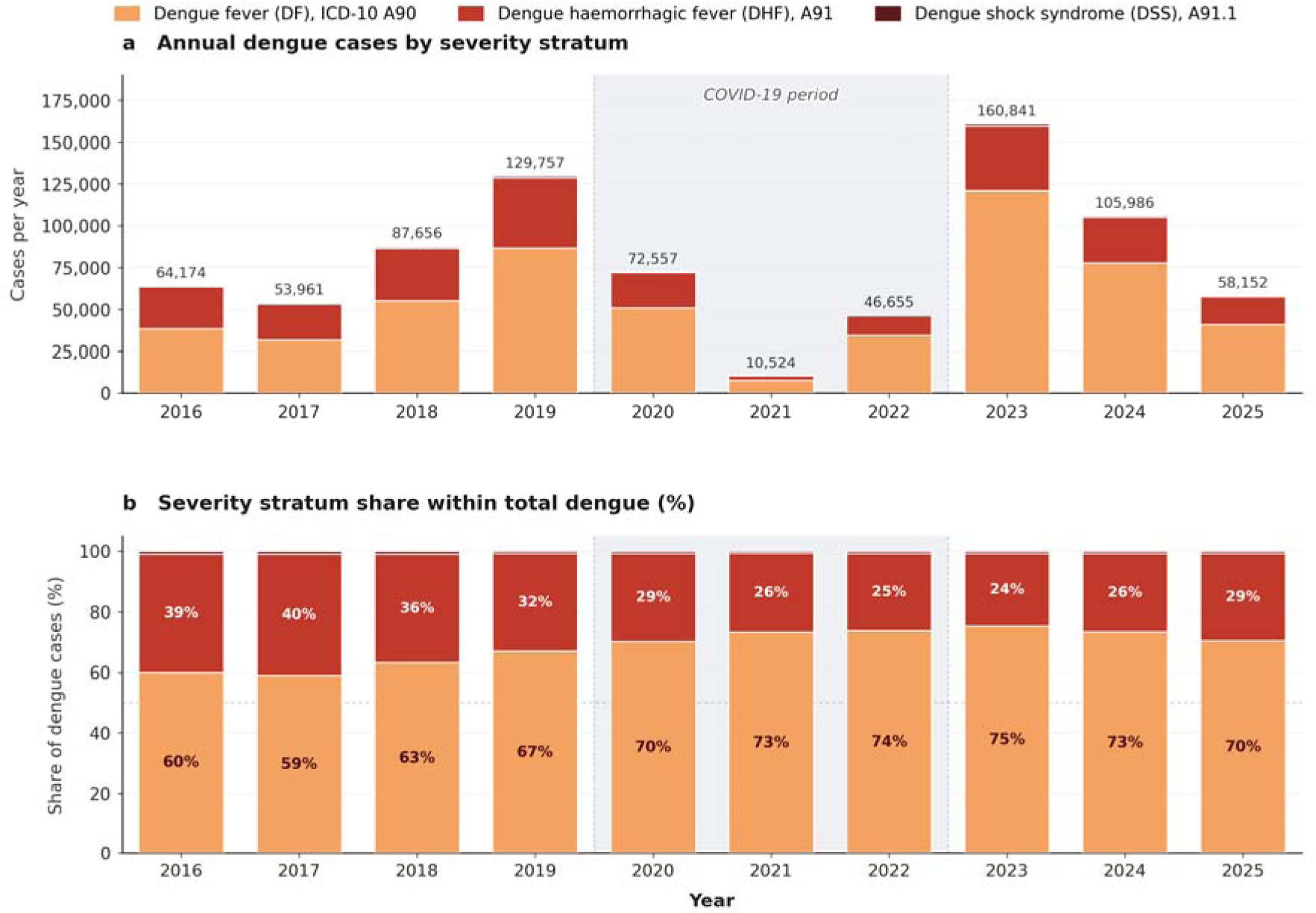
Annual dengue cases by severity stratum (a) and the share of each stratum within total dengue (b) in Thailand, 2016 to 2025. The COVID-19 period (2020 to 2022) is shaded for reference.

### Pandemic-phase shifts in national incidence

Relative to the pre-COVID phase, national incidence trajectories differed across diseases during and after the pandemic period. Dengue showed a marked decline during the pandemic, with the mean annual incidence decreasing from 126.4 per 100,000 in the pre-COVID phase to 65.4 per 100,000 (∼48.3%), followed by a rebound to 164.2 per 100,000 in the post-COVID phase (+29.8% relative to pre-COVID levels). In contrast, chikungunya demonstrated a modest increase during the pandemic period, rising from 5.8 to 6.7 per 100,000 (+16.3%), before declining substantially to 1.8 per 100,000 in the post-COVID phase (−68.1%). HFMD exhibited a pattern more similar to denguefell from 109.0 per 100,000 to 77.1 per 100,000 during the pandemic period (−29.3%), followed by recovery to 137.5 per 100,000 in the post-COVID phase (+26.1% relative to pre-COVID levels) (Table 3).

**Table 3.** Mean annual incidence by pandemic phase.

| Disease | Pre-COVID rate (2016–2019) | Pandemic rate (2020–2022) | Δ% vs Pre-COVID | Post-acute rate (2023–2025) | Δ% vs Pre-COVID |
| --- | --- | --- | --- | --- | --- |
| Dengue | 126.4 | 65.4 | –48.3% | 164.2 | +29.8% |
| Chikungunya | 5.8 | 6.7 | +16.3% | 1.8 | –68.1% |
| HFMD | 109.0 | 77.1 | –29.3% | 137.5 | +26.1% |

Note: Rates are mean annual cases per 100,000 population. Δ% = percent change in mean annual rate relative to the Pre-COVID period (2016–2019). HFMD, hand, foot, and mouth disease

### Seasonal climatology and peak timing

Mean monthly incidence by pandemic phase showed that dengue and HFMD retained their boreal-summer seasonal pattern across all three phases, whereas chikungunya exhibited a shift in seasonal timing. Dengue consistently peaked in July during the pre-COVID, pandemic, and post-COVID phases. In contrast, the peak month for chikungunya shifted from November in the pre-COVID phase to July during both the pandemic and post-acute phases; the pre-COVID November peak was primarily influenced by the late-2019 phase of the 2018–2020 nationwide outbreak. HFMD also demonstrated relatively stable seasonality, with the peak month occurring in July during the pre-COVID phase and shifted modestly to August during the pandemic and post-COVID phases (Figure 3).

**Figure 3.**
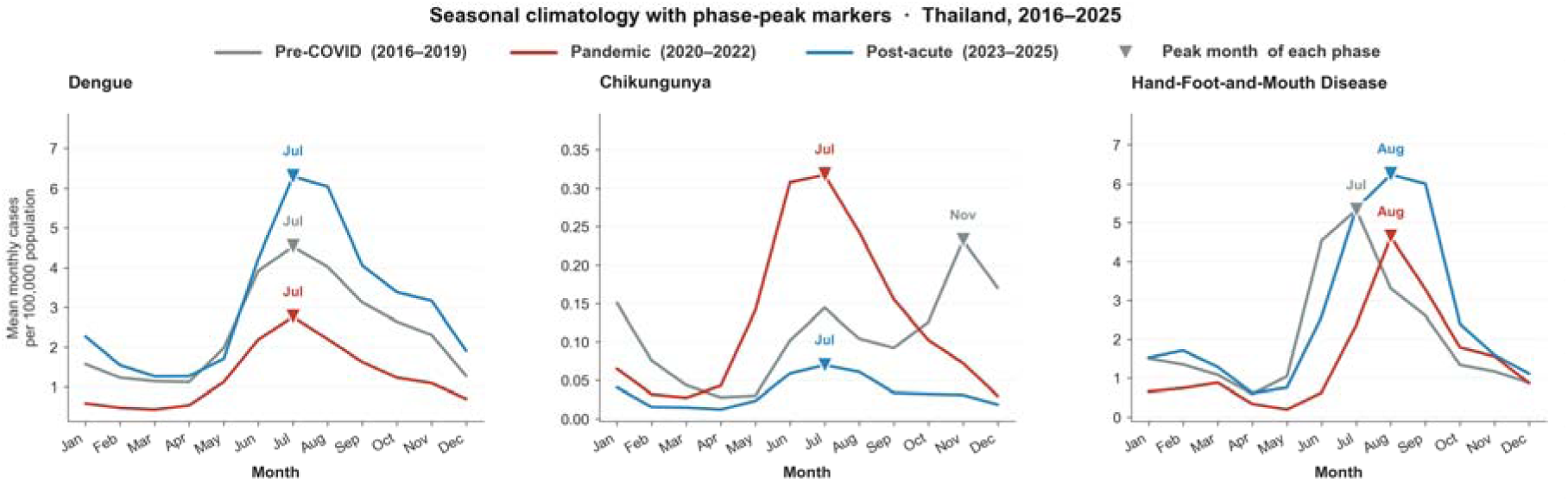
Mean monthly incidence per 100,000 population by pre-COVID, pandemic, and post-acute phases for the three diseases.

### Geographic distribution of cumulative burden

Province-level cumulative incidence over the 10-year study period demonstrated distinct spatial clustering across diseases. Dengue burden was highest in northern provinces (Mae Hong Son, Chiang Rai, Chiang Mai) and along the eastern coast (Rayong and Trat), indicating persistent high-transmission zones across both inland and coastal settings. Chikungunya incidence was concentrated along the eastern seaboard, particularly Chanthaburi, and in selected southern provinces (Songkhla and Pattani), broadly reflecting the geographic footprint of the 2018–2019 nationwide outbreak. HFMD showed the highest cumulative incidence in northern and eastern coastal provinces, with partial geographic overlap with dengue but comparatively lower burden in the deep south (Figure 4).

**Figure 4.**
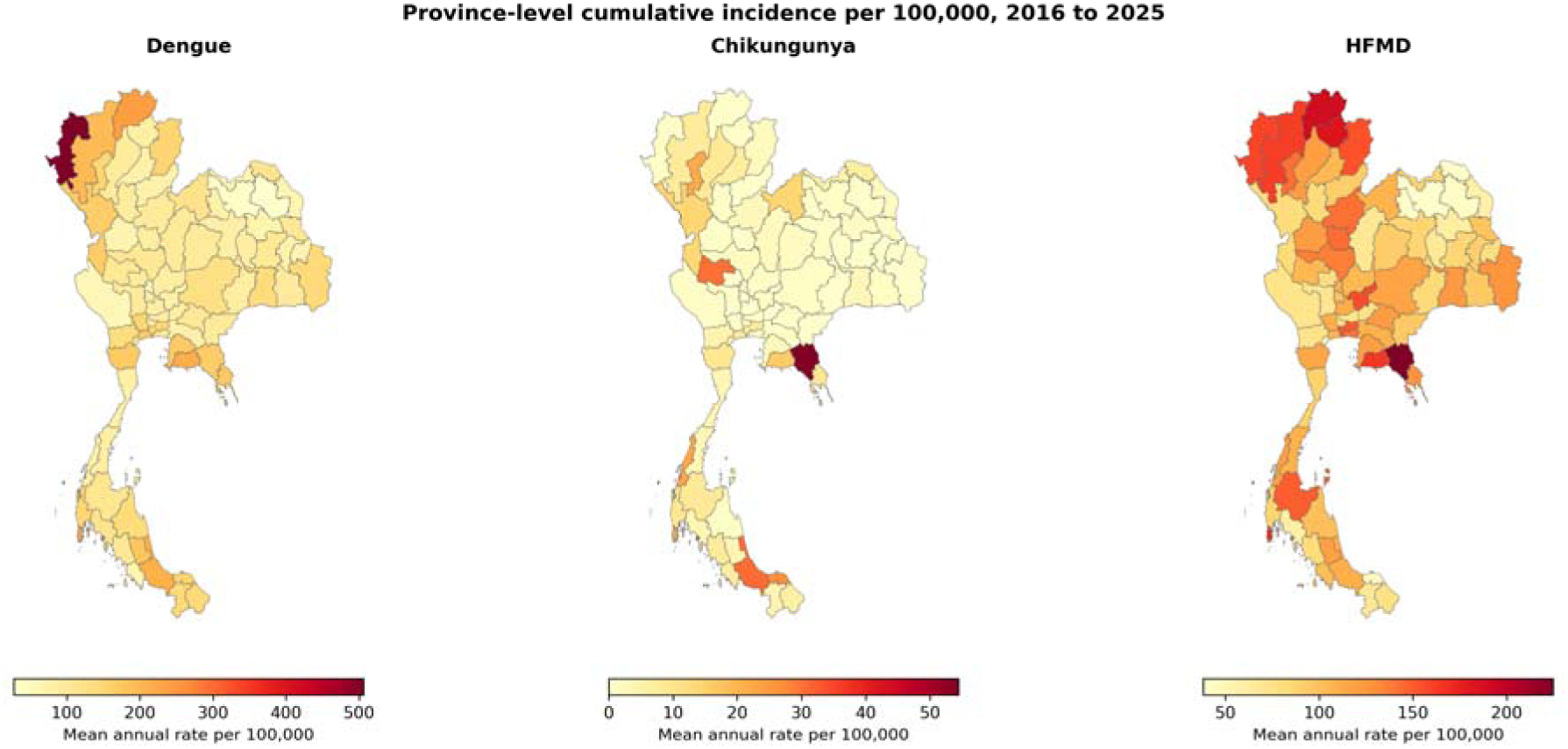
Province-level cumulative incidence per 100,000 population over 2016 to 2025 for dengue, chikungunya, and hand, foot, and mouth disease.

### Highest-burden provinces

The provinces with the highest mean annual incidence rates over 2016-2025 varied across diseases. Dengue burden was greatest in Mae Hong Son with a mean annual incidence of 504.1 per 100,000 population, followed by Chiang Rai (239.1) and Rayong (214.7). For chikungunya, Chanthaburi recorded the highest mean annual incidence (54.4 per 100,000), followed by Songkhla (30.7) and Uthai Thani (30.2). HFMD incidence was also highest in Chanthaburi (224.9 per 100,000), followed by Chiang Rai (189.6) and Phayao (182.7). Notably, Chanthaburi ranked among the five highest-burden provinces for both chikungunya and HFMD, suggesting overlapping geographic conditions that may support transmission across diseases with different epidemiological mechanisms (Table 4).

**Table 4.** Ten provinces with the highest mean annual incidence rate, by disease.

| Rank | Dengue province | Dengue rate | Chikungunya province | Chikungunya rate | HFMD province | HFMD rate |
| --- | --- | --- | --- | --- | --- | --- |
| 1 | Mae Hong Son | 504.1 | Chanthaburi | 54.4 | Chanthaburi | 224.9 |
| 2 | Chiang Rai | 239.1 | Songkhla | 30.7 | Chiang Rai | 189.6 |
| 3 | Rayong | 214.7 | Uthai Thani | 30.2 | Phayao | 182.7 |
| 4 | Phuket | 213.6 | Pattani | 27.2 | Phuket | 170.0 |
| 5 | Songkhla | 209.7 | Ranong | 22.3 | Rayong | 164.3 |
| 6 | Chiang Mai | 194.5 | Phuket | 22.0 | Chiang Mai | 161.7 |
| 7 | Phatthalung | 179.2 | Lamphun | 21.6 | Mae Hong Son | 159.3 |
| 8 | Trat | 175.3 | Rayong | 16.0 | Saraburi | 157.0 |
| 9 | Chonburi | 170.8 | Tak | 14.2 | Nan | 153.9 |
| 10 | Phetchaburi | 170.8 | Loei | 13.9 | Bangkok | 150.9 |

Note: Rates are mean annual cases per 100,000 population over 2016–2025. HFMD, hand, foot, and mouth disease

### Geographic pattern of pandemic-era change

Province-level % change in the mean annual incidence relative to the pre-COVID phase showed widespread declines during the pandemic and a partial rebound in the post-acute phase. The pandemic versus pre-COVID contrast was predominantly negative across most provinces for dengue and HFMD, whereas chikungunya showed a mixed pattern with increases in several upper-northern provinces and decreases elsewhere. The post-acute versus pre-COVID contrast was predominantly positive for dengue and HFMD across the upper north and central regions and remained predominantly negative for chikungunya nationwide (Figure 5).

**Figure 5.**
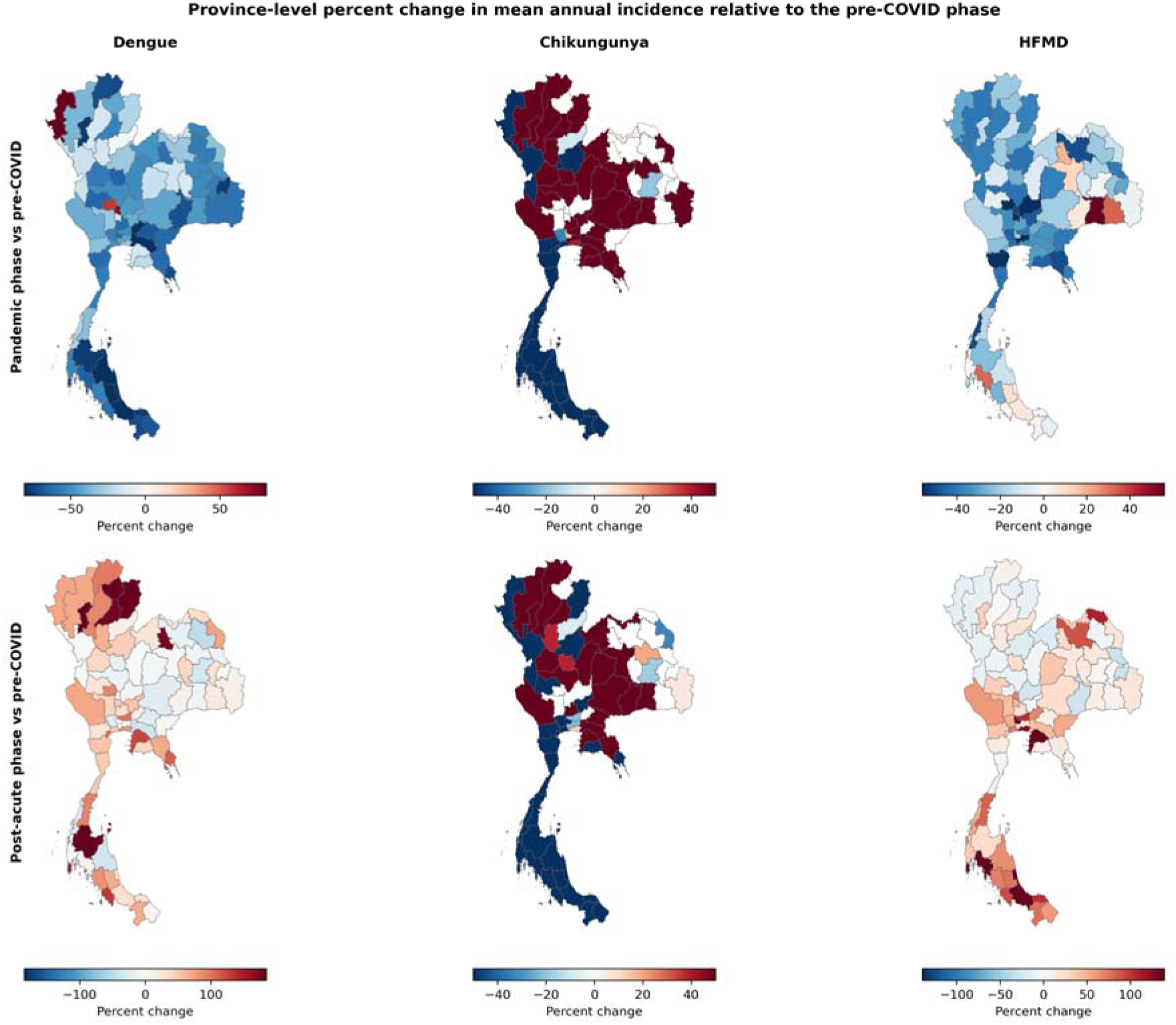
Province-level percent change in mean annual incidence during the pandemic and post-acute phases relative to the pre-COVID phase.

### Counterfactual interrupted time-series

Comparison of the fitted interrupted time-series with the no-COVID counterfactual illustrated the magnitude and direction of deviation during the pandemic and post-COVID periods for each disease. For dengue, the fitted trajectory remained below the counterfactual throughout most of the pandemic period before gradually converging toward the projected baseline during the post-COVID phase, corresponding to a cumulative difference of −9.2% relative to the counterfactual expectation. For HFMD, the observed trajectory also remained below the counterfactual during the pandemic peroid but exceeded the projected baseline in 2024-2025, resulting in an overall cumulative difference of −1.5% relative to the counterfactual. In contrast, chikungunya showed a markedly different pattern: the observed trajectory remained substantially above the seasonal baseline counterfactual derived from 2016-2017 data, with cumulative observed cases approximately 200-fold greater than the baseline expectation. This pattern reflects persistence of the 2018-2020 nationwide outbreak into the early pandemic period, rather than an isolated pandemic-associated change in transmission dynamics (Figure 6).

**Figure 6.**
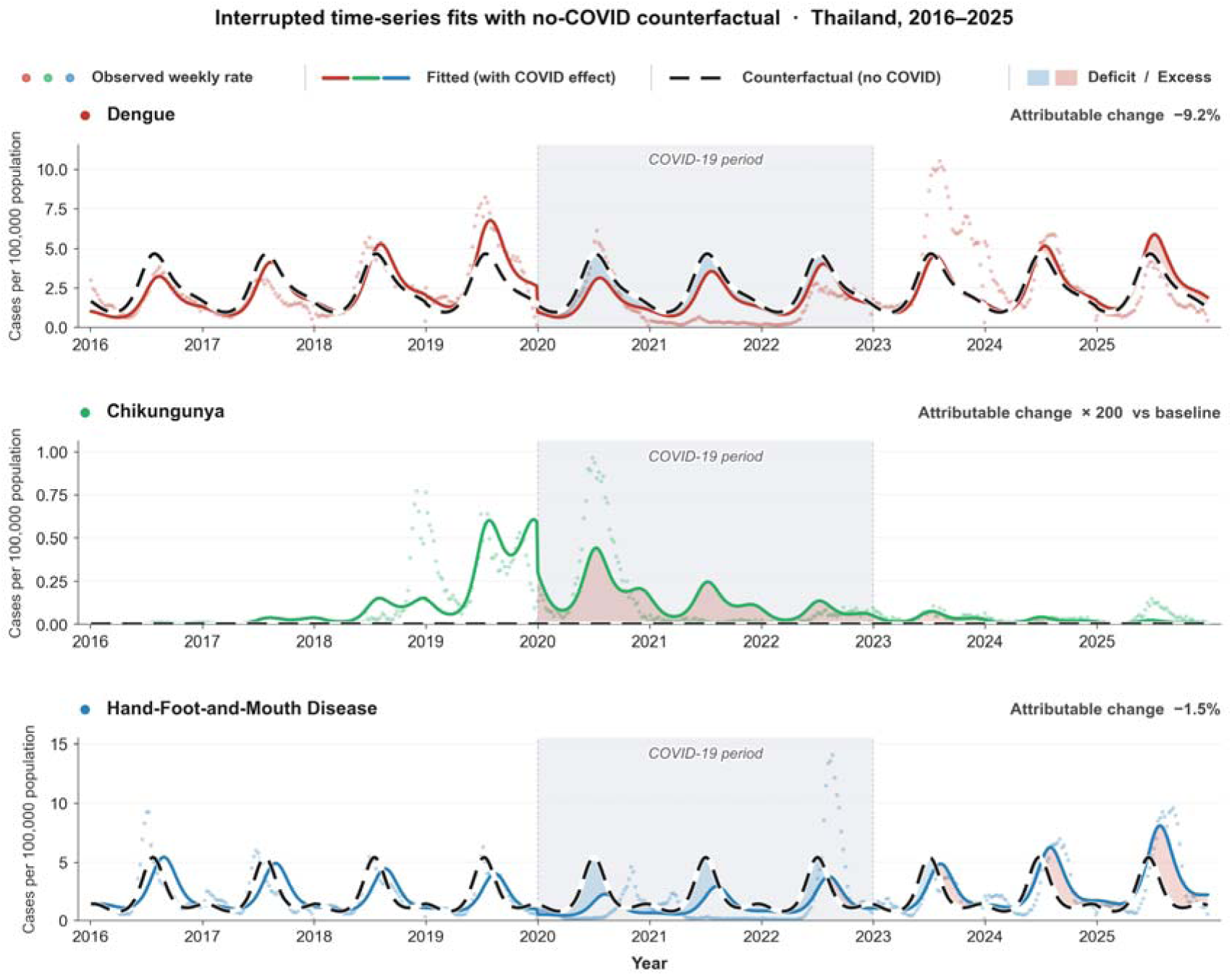
Fitted interrupted time-series and no-COVID seasonal-baseline counterfactual for each disease.

### Quantified pandemic-associated changes from interrupted time-series

Segmented regression identified statistically significant immediate level changes in incidence for all three diseases at the onset of the pandemic. At epidemiological week 1 of 2020, the estimated immediate incidence rate ratio (IRR) was 0.39 (95% CI 0.32 to 0.48; *p*<0.001) for dengue, 0.54 (0.41 to 0.71; *p*<0.001) for chikungunya, and 0.51 (0.38 to 0.69; *p*<0.001) for HFMD, corresponding to substantial downward shifts relative to the projected pre-pandemic trajectories.

Post-onset slope changes diverged across diseases. Dengue demonstrated a sustained decline of 11.4% per year (95% CI 18.2% to 3.9%; *p*=0.003). Chikungunya showed the steepest sustained decline, with an estimated annual decrease of 85.9% (88.6% to 82.6%; *p*<0.001), although interpretation should consider the influence of the preceding 2018–2020 epidemic wave. In contrast, HFMD exhibited a sustained increase of 41.9% per year (28.2% to 57.1%; *p*<0.001), indicating progressive recovery and eventual rebound following the initial pandemic-associated reduction (Table 5).

**Table 5.** Interrupted time-series estimates for the immediate level change and the sustained slope change.

| Disease | Effect | Coefficient (log-RR) | SE | Magnitude (95% CI) | P-value |
| --- | --- | --- | --- | --- | --- |
| Dengue | Immediate level change | -0.945 | 0.106 | IRR 0.39 (0.32, 0.48) | <0.001 |
|  | Sustained slope change | -0.002 | 0.001 | -11.4% per year (-18.2%, -3.9%) | 0.003 |
| Chikungunya | Immediate level change | -0.613 | 0.138 | IRR 0.54 (0.41, 0.71) | <0.001 |
|  | Sustained slope change | -0.038 | 0.002 | -85.9% per year (-88.6%, -82.6%) | <0.001 |
| HFMD | Immediate level change | -0.673 | 0.153 | IRR 0.51 (0.38, 0.69) | <0.001 |
|  | Sustained slope change | 0.007 | 0.001 | +41.9% per year (+28.2%, +57.1%) | <0.001 |

Note: Estimates from segmented quasi-Poisson regression. log-RR, log rate ratio; SE, standard error; IRR, incidence rate ratio (exponentiated coefficient); CI, confidence interval. Immediate level change is the step change at pandemic onset; sustained slope change is the per-year trend change relative to the pre-pandemic slope

### Robustness to alternative pandemic onsets

Re-fitting the interrupted time-series model using the two alternative pandemic onset definitions yielded generally consistent estimates of the immediate level change, supporting the robustness of the primary findings. For dengue, the estimated immediate IRR remained stable across definitions, with values of 0.39 at the primary onset, 0.39 at the first national lockdown, and 0.37 at the second-wave onset (*p*<0.001 for all comparisons). For chikungunya, the immediate IRR was 0.54, 0.47, and 0.05 across the three onset definitions, respectively, with all estimates remaining statistically significant. For HFMD, the corresponding IRRs were 0.51, 0.55, and 1.30; the estimate based on the second-wave onset did not reach statistical significance (*p*=0.077).

Post-onset slope estimates remained directionally consistent across onset definitions for chikungunya and HFMD. In contrast, dengue demonstrated sensitivity of the sustained slope estimate to later onset definitions, with the direction of the post-onset trend changing between the first and second alternative specifications (Supplementary Table S1). Overall, the immediate level-change estimates appeared more stable than the sustained slope estimates across alternative intervention timings.

### Cross-disease temporal coupling

Cross-correlation analysis of deseasonalised national weekly log-incidence rates demonstrateed synchronous temporal association between the two vector-borne diseases and between dengue and HFMD, whereas coupling between chikungunya and HFMD was minimal. The strongest correlation between dengue and chikungunya occurred at lag 0 (*r*=0.44), indicating that fluctuations in the two diseases tended to occur contemporaneously after adjustment for seasonal structure. Similarly, the strongest correlation between dengue and HFMD was also observed at lag 0 (*r*=0.60), suggesting partially aligned short-term temporal dynamics despite differing transmission mechanisms. In contrast, chikungunya and HFMD showed no meaningful temporal coupling; the largest observed correlation was *r*=−0.09 at lag −26 weeks, which remained within the 95% null band and was therefore consistent with no detectable association (Figure 7).

**Figure 7.**
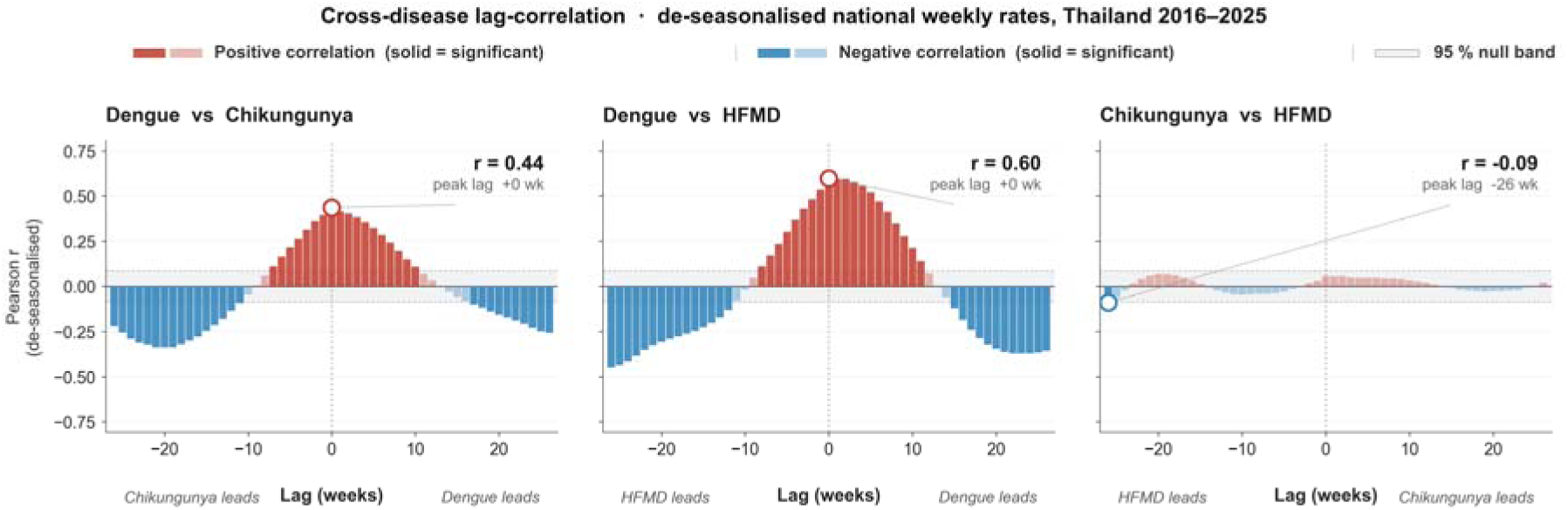
Lag correlation of de-seasonalised national weekly log-rates for each pair of diseases, with the 95 percent null band shaded.

### Differential response by transmission-mechanism

When standardized to a mechanism-specific pre-COVID weekly mean of 100, the trajectories of vector-borne and contact-mediated showed distinct temporal patterns during and after the pandemic period. The vector-borne index reached a maximum of 396 in August 2023 and declined to a minimum of 7% of the pre-COVID baseline in February 2022. In contrast, the contact-mediated index reached a higher peak of 564 in August 2022 and a lower trough of 3% baseline in March 2022, indicating a larger relative contraction followed by a more rapid rebound.

Both transmission categories exceeded their pre-pandemic baseline during the post-COVID period; however, recovery of the contact-mediated trajectory preceded recovery of the vector-borne trajectory by approximately one year (Figure 8). This temporal separation suggests that diseases with different transmission mechanisms may respond differently to large-scale disruptions in population behavior and surveillance systems.

**Figure 8.**
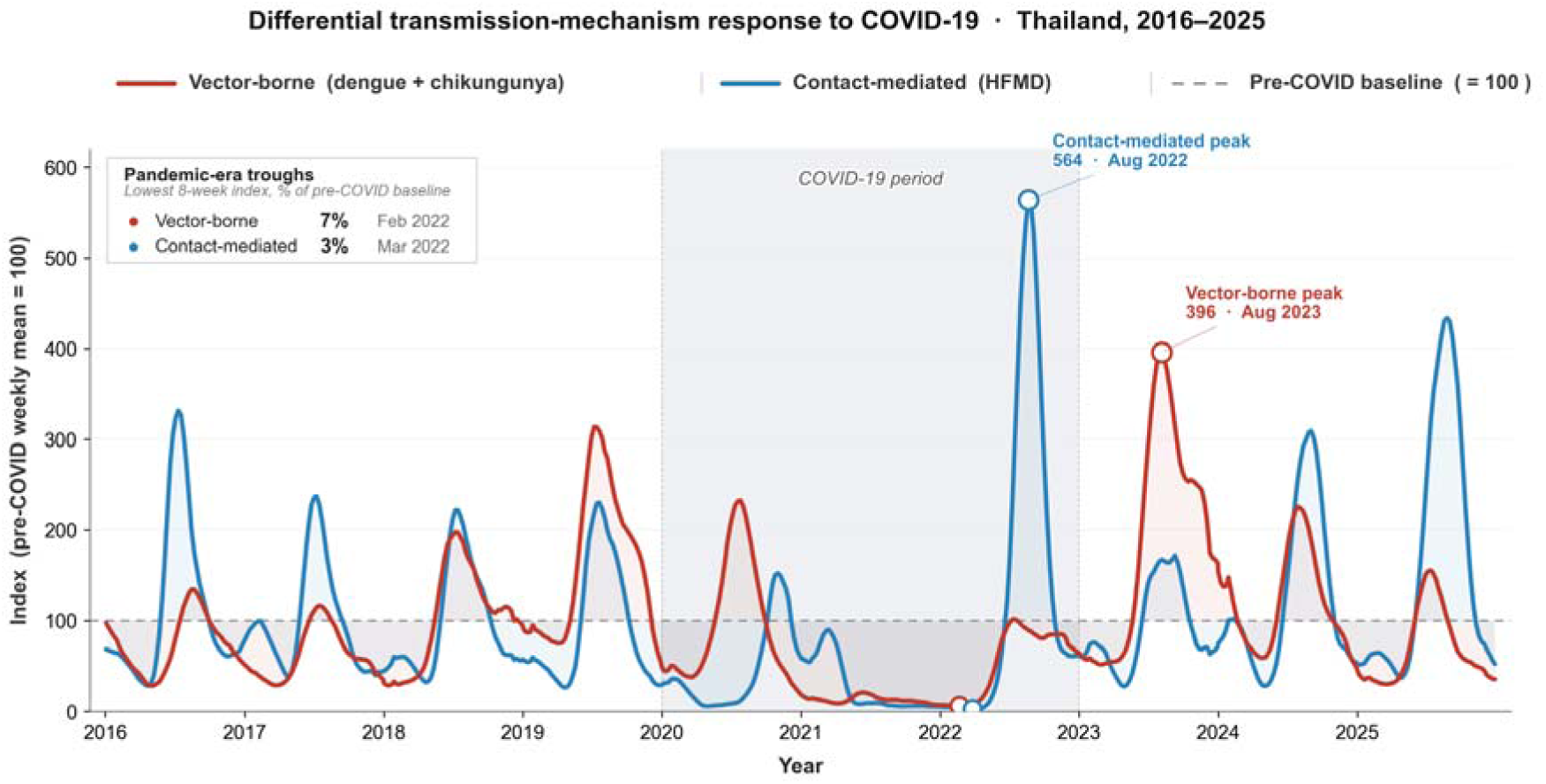
Weekly trajectories of vector-borne and contact-mediated diseases indexed to their pre-COVID weekly mean equal to 100.

## Discussion

Over a decade of nationwide notifiable disease surveillance in Thailand, dengue, chikungunya, and HFMD exhibited distinct epidemiological trajectories, with the differential response between vector-borne and contact-mediated diseases emerging as the most consistent finding. Interrupted time-series modelling estimated substantial immediate reductions in incidence at the onset of the COVID-19 pandemic across all three diseases, followed by divergent post-onset trajectories. Dengue declined sharply and subsequently rebounded above pre-pandemic levels, chikungunya entered a prolonged period of low activity following the 2018–2020 epidemic, and HFMD recovered rapidly after an initial suppression and exceeded its pre-pandemic baseline within three years. Immediate level-change estimates were broadly robust across alternative pandemic onset definitions for dengue and chikungunya, whereas the later-onset HFMD estimate did not reach statistical significance (Supplementary Table S1). Cross-disease temporal coupling further showed synchronous variation between dengue and chikungunya and between dengue and HFMD, whereas chikungunya and HFMD demonstrated no detectable temporal association.

The decadal dengue trajectory observed in Thailand is consistent with, but not identical to, the broader regional and global patterns of an acute pandemic-era trough followed by an exceptional resurgence. Recent global surveillance reports indicate that dengue incidence reached historically high levels during the post-pandemic period, with more than 10 million reported cases across 176 countries in the first seven months of 2024 alone ^16^. The Thai dataset reproduces the early pandemic dengue trough previously described in national analyses ^17^. Companion analysis of the same national surveillance dataset has further documented that the pandemic-era decline in dengue was geographically uneven, with persistent high-incidence clusters in the upper north and contracting clusters in the central region ^18^.

However, not every Southeast Asian setting followed the same trajectory; Singapore experienced a paradoxical 37% increase in dengue notifications in 2020, which was attributed to a shift in human activity into residential settings during stay-at-home measures and a corresponding rise in domestic *Aedes aegypti* exposure ^19^. This contrast suggests that pandemic-era dengue dynamics depend on a specific mix of mobility restrictions, housing structures, and pre-pandemic transmission intensity rather than lockdown stringency alone. Cluster-randomized evidence from Yogyakarta, Indonesia, has further shown that Wolbachia-based vector replacement can reduce symptomatic dengue by approximately 77% ^20^, indicating that the post-pandemic rebound observed in Thailand is not deterministic and remains amenable to vector-targeted interventions. Unlike prior single-disease analyses, our findings suggest that the magnitude and persistence of pandemic-era change may be better explained by transmission ecology than by disease identity alone.

The chikungunya trajectory observed here, in which the 2018–2019 epidemic abruptly collapsed at the start of the pandemic and has not since rebounded, is biologically plausible given both the timing of the outbreak and the genomic context of the responsible viral lineage. The 2018–2019 Thai chikungunya outbreak ^21^ was driven by a distinct sub-lineage of the Indian Ocean Lineage that had circulated in East Africa and South Asia for approximately 15 years before being introduced into Thailand from Bangladesh in 2017 ^22^. Mechanistic immunology has shown that recovery from chikungunya virus infection is associated with neutralizing antibodies and cellular responses that mediate viral clearance and resolution of acute disease ^23^. The relatively narrow seroprevalence profile generated by intense but geographically heterogeneous outbreaks, together with post-infection immunity, implies a multi-year refractory window following a major epidemic. The post-2020 quiescence in Thailand is consistent with this dynamic, combined with concurrent pandemic-era reductions in human-vector contact.

The HFMD trajectory, in which the incidence collapsed during 2020 to 2021 and then rebounded above the pre-pandemic counterfactual, is the pattern most directly aligned with the immunity debt hypothesis articulated for pediatric infections during the pandemic ^24^. A large population-based observational study from Anhui, China, attributed the early pandemic reduction in HFMD incidence to non-pharmaceutical interventions implemented for COVID-19 control ^25^, and a decade-long series from Zhejiang reported a delayed but elevated summer 2023 peak in HFMD activity following the end of the zero-COVID policy ^26^. A 2024 review of enterovirus-associated HFMD has further argued that durable global control will require sustained molecular surveillance because of the shifting balance among enterovirus A71, coxsackievirus A6, and coxsackievirus A16 ^27^. Therefore, the contemporary post-pandemic resurgence is likely to involve a serotype mix that differs from earlier epidemic cycles, reflecting the continued evolution and increased prevalence of coxsackievirus A6 documented in Thailand since the early 2010s ^28^.

The differential response between vector-borne and contact-mediated diseases observed in this study can be interpreted mechanistically by referring to the dominant transmission setting of each pathogen. Empirical evidence from Iquitos, Peru, has shown that fine-scale house-to-house human movement is the principal driver of dengue virus transmission and that mosquitoes themselves move only short distances compared with people ^29^. The global spread of *Aedes aegypti* and *Aedes albopictus* has likewise been shown to track human mobility, with future expansion projected under plausible climate trajectories ^4^, and the modelled redistribution of *Aedes*-borne virus transmission risk projects substantial poleward expansion and new northern exposures through 2080 ^30^.

In contrast, pandemic-era restrictions on inter-household mobility, mass gatherings, and school attendance had measurable, large effects on contact-mediated transmission. A multi-country analysis of approximately 1,700 non-pharmaceutical interventions across China, South Korea, Italy, Iran, France, and the United States estimated that these measures prevented or delayed approximately 61 million confirmed COVID-19 cases ^31^, and detailed work from China demonstrated that suspending intracity public transport, closing entertainment venues, and banning public gatherings reduced incidence in the early stages of the epidemic ^1^. Collectively, these studies suggest that interventions reducing population mixing disproportionately affect diseases sustained by direct human contact. Within this framework, the relatively modest and transient reduction observed for dengue in Thailand may reflect persistent peridomestic transmission, whereas the sharper decline and subsequent rebound of HFMD are consistent with the disruption and restoration of school-based mixing.

This study had several strengths. In addition to complete national coverage across all 77 provinces over a decade, the underlying harmonized surveillance dataset has been publicly deposited in a repository to support transparency, reproducibility, external validation, and future data reuse. Year-specific civil-registration population denominators were used to express incidence on a comparable rate scale throughout the study period. Furthermore, the interrupted time-series design, implemented using segmental quasi-Poisson regression with Fourier harmonics, represents an established approach for evaluating seasonally cyclical surveillance data following a single population-level eruption^12^. Sensitivity analyses at three altermative pandemic onset definitions, together with overdispersion-adjusted variance estimation, reduced dependence on arbitrary intervention timing and seasonal misspecification, consistent with the current methodological guidance for interrupted time-series studies in health services research ^32^.

Nevertheless, this study has several limitations. As a passive surveillance system, MOPH 506 reporting is shaped by clinician-to-clinician variation in case definition, laboratory throughput, and the willingness of patients to present for care, all of which were perturbed during the pandemic-era redirection of health-system resources. The analysis used aggregated reported case counts and did not adjust for serotype or genotype, which limits inferences about immune-mediated mechanisms beyond the descriptive level. Although the interrupted time-series framework is robust, it cannot definitively distinguish a true transmission interruption from a parallel attrition in surveillance sensitivity, and the introduction of a control series or a within-country sentinel benchmark is recommended for future replication ^33^. Although the within-dengue severity composition shifted toward the milder dengue fever case definition across the decade (Figure 2), the shift was monotonic and pre-dated the pandemic; therefore, it does not confound the pandemic-onset effects estimated by the ITS model. This pattern is consistent with the long-term reporting trend within the Ministry of Public Health 506 framework, the decadal-scale dengue trends reported here should be interpreted as the combined product of true incidence and this gradual reporting reclassification.

The findings are most directly generalizable to other hyperendemic Southeast Asian settings with broadly comparable climates, peridomestic *Aedes aegypti* densities, school-based mixing patterns for young children, and a centralized passive surveillance system. The mechanistic asymmetry between vector-borne and contact-mediated diseases is likely to be transportable to neighboring countries with similar transmission ecologies, although the absolute magnitude of pandemic-era change will depend on the local stringency of mobility restrictions, the duration of school closures, the timing of major pre-pandemic outbreaks, and the resilience of local laboratory capacity. Caution is warranted before extrapolating the results to subtropical or temperate settings or active sentinel surveillance systems with substantially different case-ascertainment characteristics. Because all diseases originated from the same surveillance platform, shared reporting artifacts may partially explain synchronous changes observed across diseases.

These findings have direct implications for public health practices in Thailand and Southeast Asia. The asymmetric post-pandemic recovery between vector-borne and contact-mediated diseases suggests that pandemic-era reductions should not be interpreted as durable gains, particularly for dengue, where post-pandemic resurgence is now an established global phenomenon. Wider deployment of the tetravalent dengue vaccine TAK-003, which retained 61.2% efficacy against virologically confirmed dengue and 84.1% efficacy against dengue-related hospitalization through 4.5 years of follow-up in a phase 3 trial conducted across eight dengue-endemic countries, including Thailand ^34^, together with sustained integrated vector management, is likely to be needed to limit amplification cycles. For HFMD, the pandemic-era trough and subsequent overshoot strengthen the case for routine enterovirus A71 immunization in young children where epidemiologically appropriate, given that inactivated enterovirus A71 vaccines have demonstrated greater than 90% efficacy against EV-A71-associated HFMD in phase 3 trials in China ^35^. From a surveillance standpoint, the heterogeneity in pandemic-era responses across transmission modes underscores the value of maintaining cross-disease, national-scale notifiable surveillance with year-specific population denominators throughout periods of system stress so that the differential response of communicable diseases to a future shared shock can again be quantified rather than inferred.

## Conclusion

Dengue, chikungunya, and HFMD in Thailand exhibited distinct decadal trajectories that aligned more closely with transmission ecology than with a uniform pandemic response. Although incidence declined across diseases at the onset of COVID-19, recovery patterns diverged: dengue rebounded above pre-pandemic levels, chikungunya remained suppressed following its epidemic wave, and HFMD exceeded baseline after rapid post-pandemic recovery. Immediate level-change estimates were robust across alternative pandemic onset definitions for dengue and chikungunya. These findings suggest that reductions in infectious disease incidence during periods of societal disruption should not be interpreted as durable control gains. Comparative surveillance across diseases with different transmission mechanisms may provide a more informative framework for interpreting future population-level shocks than single-disease monitoring alone.

## Abbreviations

CI: Confidence interval
COVID-19: Coronavirus disease 2019
CV-A6: Coxsackievirus A6
CV-A16: Coxsackievirus A16
DF: Dengue fever
DHF: Dengue haemorrhagic fever
DSS: Dengue shock syndrome
DOPA: Department of Provincial Administration
EV-A71: Enterovirus A71
HFMD: Hand, foot, and mouth disease
ICD-10: International Classification of Diseases, Tenth Revision
IRR: Incidence rate ratio
MOPH: Ministry of Public Health
STROBE: Strengthening the Reporting of Observational Studies in Epidemiology
TRENDS: Temporal Recent Epidemiology of Notifiable Diseases in Southeast Asia

## Declarations

## Author contributions

K. P. conceptualized and designed the study; M. M.A. performed data curation, statistical analysis, and figure generation; M. M.A. drafted the manuscript; W. P. curated the surveillance data and contributed to the epidemiological interpretation. P. and K. P. revised the manuscript; K. P. supervised the study; and all authors have read and approved the final manuscript.

## Ethics statement

The study protocol was approved as exempt by the Bamrasnaradura Infectious Diseases Institute Research Ethics Committee on 27 February 2026 (reference IRB/BIDI S002h/69_Exempt), reflecting the use of de-identified secondary surveillance data with no contact with human participants. The research was conducted in accordance with the Declaration of Helsinki. Public release of the dataset includes only aggregated, de-identified province-level records and does not contain personal identifiers or individual-level surveillance data.

## Data Availability

The aggregated province-level weekly surveillance dataset underlying this study has been publicly deposited in Zenodo under the TRENDS Dataset (2016–2025) repository. The dataset contains de-identified, aggregated surveillance records for dengue, chikungunya, and hand, foot and mouth disease, together with accompanying metadata and documentation to support reproducibility and reuse. No individual-level data are included. Dataset DOI: <u>10.5281/zenodo.20269896</u>.

## Clinical trial registration

Not applicable.

## Consent to publish

Not applicable.

## Consent for publication

Not applicable.

## Acknowledgements

The authors acknowledge the Department of Disease Control (DDC), Ministry of Public Health, Thailand, for maintaining the national notifiable disease surveillance system and supporting access to the aggregated surveillance data used in this study. We also thank surveillance personnel, public health officers, and healthcare providers across all provinces whose routine reporting enabled the development and public dissemination of the TRENDS-THAI dataset.

## Generative AI Use Statement

The authors used Paperpal AI solely for language editing and improvement of manuscript clarity and readability. No generative artificial intelligence tools were used for study design, data collection, data analysis, interpretation of findings, or generation of scientific conclusions. All scientific content was reviewed and verified by the authors, who take full responsibility for the accuracy and integrity of the work.

## References

1. Tian H, Liu Y, Li Y, et al. An investigation of transmission control measures during the first 50 days of the COVID-19 epidemic in China. Science 2020;368(6491):638–42. doi: 10.1126/science.abb6105

2. Olsen SJ, Azziz-Baumgartner E, Budd AP, et al. Decreased Influenza Activity During the COVID-19 Pandemic — United States, Australia, Chile, and South Africa, 2020. MMWR Morb Mortal Wkly Rep 2020;69(37):1305–09. doi: 10.15585/mmwr.mm6937a6

3. Roberton T, Carter ED, Chou VB, et al. Early estimates of the indirect effects of the COVID-19 pandemic on maternal and child mortality in low-income and middle-income countries: a modelling study. The Lancet Global Health 2020;8(7):e901–e08. doi: 10.1016/S2214-109X(20)30229-1

4. Kraemer MUG, Reiner RC, Brady OJ, et al. Past and future spread of the arbovirus vectors Aedes aegypti and Aedes albopictus. Nat Microbiol 2019;4(5):854–63. doi: 10.1038/s41564-019-0376-y

5. Du M, Jing W, Liu M, et al. The Global Trends and Regional Differences in Incidence of Dengue Infection from 1990 to 2019: An Analysis from the Global Burden of Disease Study 2019. Infect Dis Ther 2021;10(3):1625–43. doi: 10.1007/s40121-021-00470-2

6. Brady OJ, Hay SI. The Global Expansion of Dengue: How Aedes aegypti Mosquitoes Enabled the First Pandemic Arbovirus. Annu Rev Entomol 2020;65(1):191–208. doi: 10.1146/annurev-ento-011019-024918

7. Cowling BJ, Ali ST, Ng TWY, et al. Impact assessment of non-pharmaceutical interventions against coronavirus disease 2019 and influenza in Hong Kong: an observational study. The Lancet Public Health 2020;5(5):e279–e88. doi: 10.1016/S2468-2667(20)30090-6

8. Phadungsombat J, Imad H, Rahman M, et al. A Novel Sub-Lineage of Chikungunya Virus East/Central/South African Genotype Indian Ocean Lineage Caused Sequential Outbreaks in Bangladesh and Thailand. Viruses 2020;12(11):1319. doi: 10.3390/v12111319

9. Xing W, Liao Q, Viboud C, et al. Hand, foot, and mouth disease in China, 2008–12: an epidemiological study. The Lancet Infectious Diseases 2014;14(4):308–18. doi: 10.1016/S1473-3099(13)70342-6

10. Puenpa J, Wanlapakorn N, Vongpunsawad S, et al. The History of Enterovirus A71 Outbreaks and Molecular Epidemiology in the Asia-Pacific Region. J Biomed Sci 2019;26(1):75. doi: 10.1186/s12929-019-0573-2

11. Huang AT, Takahashi S, Salje H, et al. Assessing the role of multiple mechanisms increasing the age of dengue cases in Thailand. Proc Natl Acad Sci USA 2022;119(20):e2115790119. doi: 10.1073/pnas.2115790119

12. Wagner AK, Soumerai SB, Zhang F, et al. Segmented regression analysis of interrupted time series studies in medication use research. J Clin Pharm Ther 2002;27(4):299–309. doi: 10.1046/j.1365-2710.2002.00430.x

13. Elm Ev, Altman DG, Egger M, et al. Strengthening the reporting of observational studies in epidemiology (STROBE) statement: guidelines for reporting observational studies. BMJ 2007;335(7624):806–08. doi: 10.1136/bmj.39335.541782.AD

14. Pongpirul WA, Mohamed Mustaf; Pongpirul, Krit. TRENDS: Temporal Recent Epidemiology of Notifiable Diseases in Southeast Asia. In: Bamrasnaradura Infectious Diseases Institute DoDC, Ministry of Public Health, Thailand, Center of Excellence in Preventive and Integrative Medicine (CE-PIM) and Department of Preventive and Social Medicine, Faculty of Medicine, Chulalongkorn University, ed. v1 ed: Zenodo, 2026.

15. Lopez Bernal J, Cummins S, Gasparrini A. Interrupted time series regression for the evaluation of public health interventions: a tutorial. International Journal of Epidemiology 2016:dyw098. doi: 10.1093/ije/dyw098

16. The L. Dengue: the threat to health now and in the future. The Lancet 2024;404(10450):311. doi: 10.1016/S0140-6736(24)01542-3

17. Tangsathapornpong A, Thisyakorn U. Dengue amid COVID-19 pandemic. PLOS Glob Public Health 2023;3(2):e0001558. doi: 10.1371/journal.pgph.0001558

18. Saita S, Maeakhian S, Silawan T. Temporal Variations and Spatial Clusters of Dengue in Thailand: Longitudinal Study before and during the Coronavirus Disease (COVID-19) Pandemic. TropicalMed 2022;7(8):171. doi: 10.3390/tropicalmed7080171

19. Lim JT, Chew LZX, Choo ELW, et al. Increased Dengue Transmissions in Singapore Attributable to SARS-CoV-2 Social Distancing Measures. The Journal of Infectious Diseases 2020;223(3):399–402. doi: 10.1093/infdis/jiaa619

20. Utarini A, Indriani C, Ahmad RA, et al. Efficacy of Wolbachia-Infected Mosquito Deployments for the Control of Dengue. N Engl J Med 2021;384(23):2177–86. doi: 10.1056/NEJMoa2030243

21. Khongwichit S, Chansaenroj J, Thongmee T, et al. Large-scale outbreak of Chikungunya virus infection in Thailand, 2018–2019. PLoS ONE 2021;16(3):e0247314. doi: 10.1371/journal.pone.0247314

22. Krambrich J, Mihalič F, Gaunt MW, et al. The evolutionary and molecular history of a chikungunya virus outbreak lineage. PLoS Negl Trop Dis 2024;18(7):e0012349. doi: 10.1371/journal.pntd.0012349

23. Srivastava P, Kumar A, Hasan A, et al. Disease Resolution in Chikungunya—What Decides the Outcome? Front Immunol 2020;11 doi: 10.3389/fimmu.2020.00695

24. Cohen R, Ashman M, Taha M-K, et al. Pediatric Infectious Disease Group (GPIP) position paper on the immune debt of the COVID-19 pandemic in childhood, how can we fill the immunity gap? Infectious Diseases Now 2021;51(5):418–23. doi: 10.1016/j.idnow.2021.05.004

25. Ma W, Li X, Wang N, et al. Impact of non-pharmacological interventions on incidence of hand, foot and mouth disease during the COVID-19 pandemic: a large population-based observational study. BMC Infect Dis 2024;24(1) doi: 10.1186/s12879-024-10252-z

26. Ding Z, Lu Q, Wu H, et al. Trend of hand, foot and mouth disease before, during, and after China’s COVID control policies in Zhejiang, China. Front Public Health 2024;12 doi: 10.3389/fpubh.2024.1472944

27. Huang C-Y, Su S-B, Chen K-T. A review of enterovirus-associated hand-foot and mouth disease: preventive strategies and the need for a global enterovirus surveillance network. Pathogens and Global Health 2024;118(7-8):538–48. doi: 10.1080/20477724.2024.2400424

28. Noisumdaeng P, Puthavathana P. Molecular evolutionary dynamics of enterovirus A71, coxsackievirus A16 and coxsackievirus A6 causing hand, foot and mouth disease in Thailand, 2000–2022. Sci Rep 2023;13(1) doi: 10.1038/s41598-023-44644-z

29. Stoddard ST, Forshey BM, Morrison AC, et al. House-to-house human movement drives dengue virus transmission. Proc Natl Acad Sci USA 2012;110(3):994–99. doi: 10.1073/pnas.1213349110

30. Ryan SJ, Carlson CJ, Mordecai EA, et al. Global expansion and redistribution of Aedes-borne virus transmission risk with climate change. PLoS Negl Trop Dis 2019;13(3):e0007213. doi: 10.1371/journal.pntd.0007213

31. Hsiang S, Allen D, Annan-Phan S, et al. The effect of large-scale anti-contagion policies on the COVID-19 pandemic. Nature 2020;584(7820):262–67. doi: 10.1038/s41586-020-2404-8

32. Penfold RB, Zhang F. Use of Interrupted Time Series Analysis in Evaluating Health Care Quality Improvements. Academic Pediatrics 2013;13(6):S38–S44. doi: 10.1016/j.acap.2013.08.002

33. Lopez Bernal J, Cummins S, Gasparrini A. The use of controls in interrupted time series studies of public health interventions. International Journal of Epidemiology 2018;47(6):2082–93. doi: 10.1093/ije/dyy135

34. Tricou V, Yu D, Reynales H, et al. Long-term efficacy and safety of a tetravalent dengue vaccine (TAK-003): 4·5-year results from a phase 3, randomised, double-blind, placebo-controlled trial. The Lancet Global Health 2024;12(2):e257–e70. doi: 10.1016/S2214-109X(23)00522-3

35. Zhu F, Xu W, Xia J, et al. Efficacy, Safety, and Immunogenicity of an Enterovirus 71 Vaccine in China. N Engl J Med 2014;370(9):818–28. doi: 10.1056/NEJMoa1304923

