## Supplementary Table S1 for "TRENDS-Thai: decadal trends of dengue, chikungunya, and hand, foot, and mouth disease in Thailand (2016–2025): a multi-disease time-series analysis of COVID-19 disruption"

**Table S1.** Sensitivity analysis: interrupted time-series estimates at three pandemic onset definitions

| Disease | Onset | Onset date | Level IRR (95% CI) | Level p | Slope per year (95% CI) | Slope p |
| --- | --- | --- | --- | --- | --- | --- |
| Dengue | Primary | 2020-01-01 | 0.39 (0.32, 0.48) | <0.001 | -11.4% (-18.2%, -3.9%) | 0.003 |
|  | Sensitivity analysis 1 | 2020-03-22 | 0.39 (0.32, 0.49) | <0.001 | -8.5% (-15.4%, -0.9%) | 0.028 |
|  | Sensitivity analysis 2 | 2020-10-12 | 0.37 (0.30, 0.47) | <0.001 | +8.2% (+0.5%, +16.5%) | 0.037 |
| Chikungunya | Primary | 2020-01-01 | 0.54 (0.41, 0.71) | <0.001 | -85.9% (-88.6%, -82.6%) | <0.001 |
|  | Sensitivity analysis 1 | 2020-03-22 | 0.47 (0.35, 0.62) | <0.001 | -84.2% (-87.1%, -80.6%) | <0.001 |
|  | Sensitivity analysis 2 | 2020-10-12 | 0.05 (0.04, 0.07) | <0.001 | -63.0% (-67.7%, -57.7%) | <0.001 |
| HFMD | Primary | 2020-01-01 | 0.51 (0.38, 0.69) | <0.001 | +41.9% (+28.2%, +57.1%) | <0.001 |
|  | Sensitivity analysis 1 | 2020-03-22 | 0.55 (0.41, 0.75) | <0.001 | +44.1% (+30.4%, +59.2%) | <0.001 |
|  | Sensitivity analysis 2 | 2020-10-12 | 1.30 (0.97, 1.75) | 0.077 | +52.0% (+38.9%, +66.4%) | <0.001 |

Note. IRR, incidence rate ratio; CI, confidence interval. Primary onset 2020-01-01 (analytical baseline); Sensitivity analysis 1 = 2020-03-22 (week 12 of 2020, first national lockdown); Sensitivity analysis 2 = 2020-10-12 (week 42 of 2020, second wave).
